# Pathogenic variation in insulin resistance genes is common in polycystic ovary syndrome (PCOS): a strategy for causal gene discovery using whole-exome sequencing (WES) in complex traits

**DOI:** 10.1101/2025.08.13.25333592

**Authors:** Rosemary Bauer, Chloe Parker, Zulma Cardona Matos, M. Geoffrey Hayes, Allen R. Kunselman, Richard S. Legro, Corrine K. Welt, Margrit Urbanek

## Abstract

Polycystic ovary syndrome (PCOS) is a complex, multi-system, heritable endocrinopathy that is a common cause of anovulatory infertility in reproductive-aged women. While insulin resistance (IR) is not a diagnostic feature, it is widespread in women with PCOS, and often more severe than in women of similar age and BMI. Conversely, women with rare Mendelian disorders of IR also present with features of PCOS. We hypothesize that PCOS is driven by underlying IR, which can be evaluated through a genetic approach. We curated and stratified 310 genes related to three mechanisms of IR using molecular and clinical criteria. We evaluated protein-altering genetic variation in 102 insulin signaling genes, 29 obesity genes, and 22 dyslipidemia genes from whole-exome sequencing data from 675 PCOS patients. 40 insulin signaling genes, 12 obesity genes, and 10 dyslipidemia genes were significantly enriched for protein-altering variation in PCOS cases compared to healthy population controls. Variants in these 62 significantly enriched genes affected 51% of PCOS cases in our study cohort. The 15 highest ranked genes were selected for follow-up: *LMNA, LEPR, KCNJ11, BSCL2, ACACA, NTRK2, GCK, ABCC8, SLC2A2, POMC, MC4R, TBC1D4, INSR, NR0B2,* and *GCKR*. 50% of variants identified in these 15 genes were pathogenic, 35% were likely pathogenic, and only 15% were variants of uncertain significance. These findings support IR as a central pathway in PCOS. Furthermore, this study demonstrates that a candidate pathway approach with sufficient pre-processing can successfully identify functionally relevant variants and genes underlying complex traits.

## Introduction

Polycystic ovary syndrome (PCOS) (MIM #184700) is a common, complex, multi-system endocrinopathy and the leading cause of anovulatory infertility in reproductive-aged women (1–3). The defining diagnostic criteria are hyperandrogenemia, irregular menses, and polycystic ovarian morphology. In addition to these cardinal features, PCOS patients exhibit metabolic impairments such as high rates of obesity, dyslipidemia, insulin resistance (IR), and type 2 diabetes (T2D) (4). Furthermore, IR in women with PCOS is more severe than in age- and BMI-matched women without PCOS (4). PCOS has a strong genetic component as established by twin studies and pedigree analyses (5, 6). Genome-wide association studies (GWAS) have revealed numerous candidate loci (7, 8); however, these only account for a small portion of the heritability. Thus, rare, high-effect variants may contribute to the missing heritability (9).

Genetic studies of complex traits are expanding their investigations beyond GWAS towards comprehensive sequencing technologies such as whole-exome sequencing (WES) to capture rare coding variants that may underlie disease etiology. Examples include Alzheimer’s disease (reviewed in (10)), chronic obstructive pulmonary disease (COPD) (reviewed in (11)), and endometriosis (reviewed in (12)). Notably, some complex diseases have related Mendelian, more severe forms. Highly penetrant, monogenic forms of a trait with well-characterized heritability can serve as informative models for the overarching complex trait. Such examples include monogenic familial hypercholesterolemia within the broader trait of high cholesterol levels (reviewed in (13)) and monogenic maturity-onset diabetes of the young (MODY) in relation to T2D (reviewed in (14)). Like these examples, PCOS may similarly encompass rare, monogenic forms of disease that contribute to its genetic and phenotypic heterogeneity. Genetic sequencing studies by our lab and others have identified several prominent genes with rare, high-effect variants accounting for a small portion of heritability (15–18). Importantly, we recently identified rare variants in *LMNA* significantly associated with PCOS cases in two independent cohorts (16). Rare pathogenic variants in *LMNA* are associated with a number of pathologies, but most relevant to PCOS is Dunnigan’s Familial Partial Lipodystrophy (FPLD2) (MIM #151660), a disorder of lipid storage and severe IR that also presents with hyperandrogenemia and irregular menses in female patients (19, 20). Therefore, we hypothesize that within the pool of patients presenting with the features of PCOS, there exists a portion of women who have underlying monogenic disorders, which could include other disorders of IR. Identifying these patients can aid in their targeted treatment and inform the overall genetic architecture of PCOS.

With the advent of sequencing technologies including WES, generation of sequence data has become increasingly accessible and rapid; however, the ability to interpret findings from this growing amount of genetic data remains a challenge, especially outside the context of Mendelian disease (21). This contributes to the accumulation of variants of uncertain significance (VUS) which have limited diagnostic or clinical utility (22). Because of this phenomenon, our efforts to apply WES to identify rare, monogenic contributors to PCOS requires substantial pre-processing of genes to ensure that the variants we identify are interpretable and relevant to the trait.

Given the phenotypic overlap between IR and PCOS, investigating the genetic architecture of IR may reveal novel genes relevant to PCOS etiology. IR is a complex condition attributable to multiple biological systems. Molecular mechanisms resulting in IR include direct impairment of the insulin signaling pathway, indirect effect through either impaired energy homeostasis resulting in obesity, or impaired adipose storage resulting in dyslipidemia (23, 24). Numerous genetic causes of severe IR have been identified which aid in these categorizations (24). While several studies have examined common GWAS variants associated with these pathologies in the context of PCOS (25–29), few have investigated IR genes for coding variants using WES. The most comprehensive analysis to date of IR genes in the context of PCOS was completed in a small cohort pre-selected for IR (15). The present study is the first to do so in a comprehensive, unbiased manner in a cohort not pre-selected for any metabolic features.

To test the hypothesis that genetic forms of IR contribute to the etiology of PCOS, we first generated a comprehensive list of 310 genes related to three classes of genetic IR: insulin signaling, obesity, and dyslipidemia. We then systematically prioritized the likely impact of these genes on IR etiology using molecular, clinical, and statistical data. Genes with sufficient *a priori* evidence were assayed for protein-altering variants in a cohort of 675 women with PCOS (Figure 1). Finally, we selected 15 PCOS candidate genes with the strongest evidence for a role in PCOS pathogenesis for genetic analyses at the variant level.

**Figure 1.**
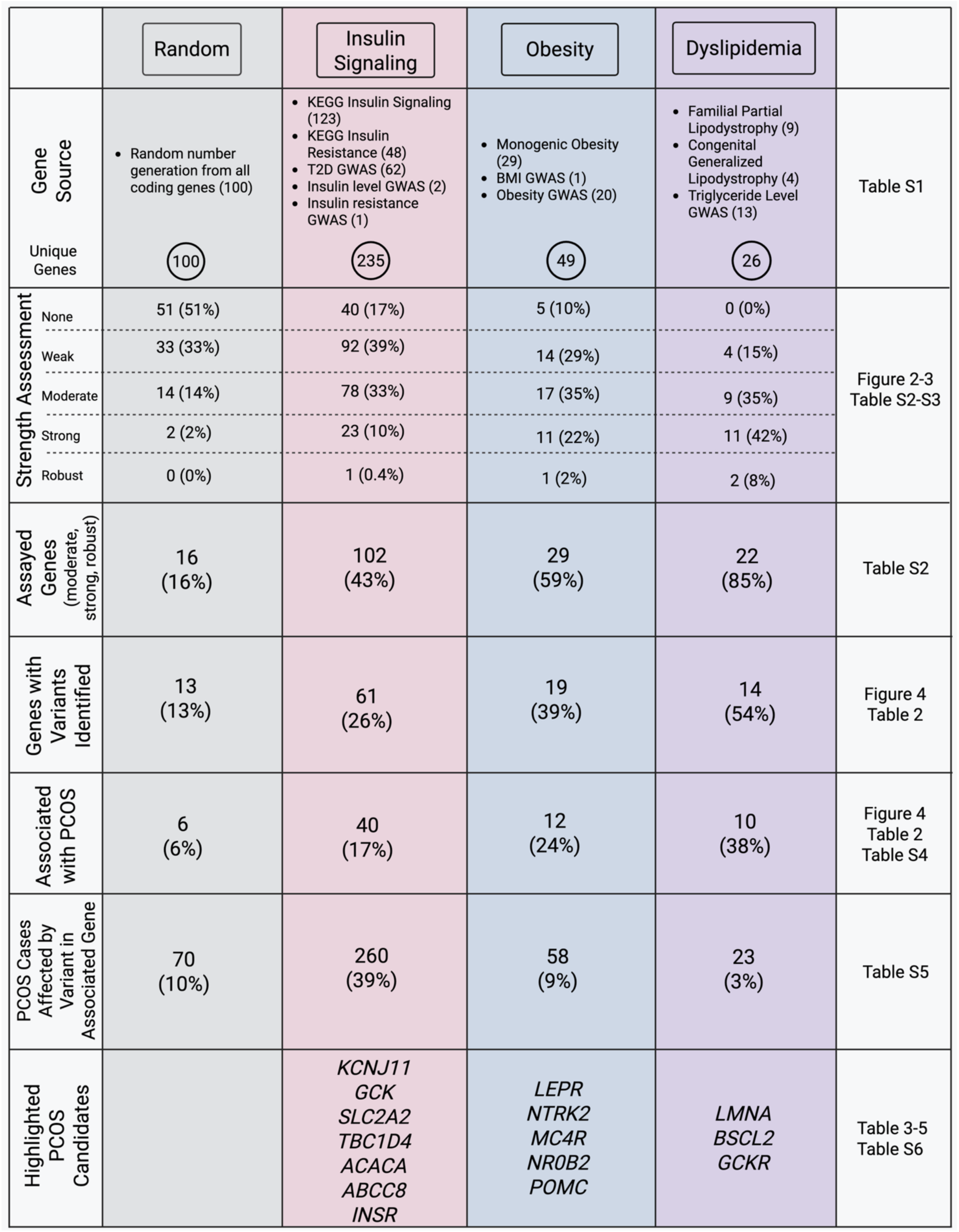
Summary of gene selection, prioritization, and association with PCOS. Percentages represent positive genes relative to total unique genes included in each gene set, except for the PCOS cases affected by a variant, which represents the number of PCOS cases affected by a variant relative to total PCOS cases (n=675).

## Subjects and Methods

### Subjects

Our study includes subjects from two PCOS cohort studies. The first cohort was recruited for phenotypic and genetic studies of PCOS and was approved by the IRB of Massachusetts General Hospital (MGH) and University of Utah. All subjects were self-reported White of European ancestry, ages 18-45 years old and in good general health. Their inclusion and exclusion criteria have been described (30, 31). PCOS cases satisfied NIH criteria, requiring ≤9 menses per year and biochemical or clinical evidence of hyperandrogenism (30, 31).

Normoandrogenic, regularly cycling controls were recruited and phenotyped alongside PCOS cases. 376 PCOS cases and all 235 control subjects originate from this study. The remaining 299 PCOS cases in our study are participants of the Pregnancy in Polycystic Ovary Syndrome II (PPCOSII) Trial recruited in Hershey, PA (32, 33). PCOS cases in PPCOSII fulfill a modified Rotterdam criteria (34), requiring ovulatory dysfunction and either hyperandrogenism or polycystic ovarian morphology (32, 33). All PCOS subjects from PPCOSII are of self-reported European ancestry. All participants provided informed consent for genetic studies.

### Hormone and Biochemical Assays

Hormones, lipids, glucose, and insulin were measured from blood samples as previously described (30, 31, 33). Comparisons of variables were corrected for study cohort as a proxy for assay methodology.

### Statistical Analysis

Statistical tests were computed in GraphPad Prism (version 10.0.3 for macOS, GraphPad Software, Boston, MA www.graphpad.com) and R Studio (version 2022.12.0.353). To compare cases and controls, each variable was modeled in a linear model assessing the contributions of age, BMI, case/control status, and study site. Homogeneity of residuals in the linear model were evaluated using the olsrr package in R. Variables with non-normal residuals were transformed to achieve normality.

To assess enrichment of variants in PCOS cases, PCOS case frequency was compared to gnomAD v2.1.1 non-Finish European population controls. Odds ratios (OR) and Fisher’s exact tests were computed as a gene-based burden. The 95% CI of the OR was computed by the Woolf logit interval. A 95% CI excluding 1 was considered significant. Fisher’s exact test was considered nominally significant if p<0.05, considered study-wide (308 genes) significant if p<1.6×10^-4^, and considered significant at the exome-wide (20,000 genes) level if p<2.5×10^-6^.

### Exome Sequencing

Genomic DNA extracted from peripheral blood was sequenced by WES at CIDR as described previously (17). Library preparation and enrichment were prepared using a low-input library prep protocol developed at CIDR. DNA was processed and amplified using Roche’s Kapa Hyper prep kit and Kapa HiFi enzyme kit. Sequencing was performed on the Illumina NovaSeq 6000 platform using 100 base pair paired end runs using Illumina NovaSeq 6000 S4 Reagent Kit and NovaSeq Xp 4-Lane kit workflow.

Alignment to reference genome build 38 and variant calling were completed using the CIDRSeqSuite pipeline (17). Quality control was performed at CIDR using Illumina InfiniumQCArray-24v1-0 array to confirm sex and ancestry, and to identify DNA sample contamination, unexpected duplicates, or relatedness. Annotation of called SNVs and indels was executed using SnpEff and SnpSift (35, 36). The Genome Aggregation Database (gnomAD) v3.1.2 (37) MAF was ascertained. Annotated variants were filtered based on Phred quality score ≥ 20, read depth ≥ 25,000, mapping quality ≥ 58.75 and ≤ 61.25, variant quality score log odds ≥ 7.81, exonic-region, variant type (predicted missense, nonsense, frameshift, and splice site variants), and Rare Exome Variant Ensemble Learner (REVEL) (38) score ≥ 0.6.

### Gene Selection

Four sets of genes were identified for prioritization in this study. First, “100 random genes” were identified using a random number generation to generate 100 numbers between 1-22394, corresponding to line numbers in a spreadsheet of all RefSeq Select genes (genome.ucsc.edu). The remaining gene sets refer to the three molecular mechanisms of IR investigated in this study: insulin signaling, obesity, and dyslipidemia. All three gene sets were identified by database or literature search and through known GWAS associations. The GWAS Catalog (ebi.ac.uk/gwas) was queried on 8/12/2024 to identify GWAS loci. Loci were required to have one “qualifying” GWAS association that fulfilled any of the following strength-based criteria: Z score increase or decrease by ≥ 3, ≥ 10% change in measurement (i.e. insulin or triglycerides), or OR > 1.20. Finally, identified loci were evaluated for eQTL or disease association in the Online Mendelian Inheritance in Man (39) (OMIM) (omim.org) database ensuring a strong relationship between the GWAS annotated gene and a phenotype. GWAS-associated genes needed to fulfil both a strength-based criterion (Z ≥ ±3, ≥ 10% change, or OR > 1.20) and have an OMIM trait association to be evaluated further. To identify the gene set known as “insulin signaling,” the Kyoto Encyclopedia of Genes and Genomes (KEGG) (genome.jp/kegg) was queried for two KEGG pathways on 4/11/2024: first “insulin signaling pathway” and then “insulin resistance.” Genes in the KEGG IR gene set were only added if they were unique and did not duplicate genes from the KEGG insulin signaling gene set. Qualifying GWAS loci associated with three traits were added to the insulin signaling gene set: T2D, IR, and insulin level. The “obesity” and “dyslipidemia” gene sets were identified through literature search in July 2024. Qualifying GWAS loci associated with BMI and obesity were included in the obesity gene set, and GWAS loci for triglyceride levels were included in the dyslipidemia gene set.

### Gene Prioritization

Genes were evaluated for their weighted candidate gene confidence level using a 4-point system (Figure 2). One point was awarded for a high degree of evolutionary constraint on missense variation (37), defined as Z > 3 in the Genome Aggregation Database (40) (gnomAD) (gnomad.broadinstitute.org) v4 on 8/9/2024. One point was awarded for any OMIM gene-phenotype relationship queried on 8/9/2024. An additional point was possible if the gene-phenotype relationship in OMIM was an IR-related phenotype, or had features of metabolic impairment, glucose dysregulation, hyperinsulinemia, hyperphagia, obesity, dyslipidemia, menstrual dysfunction, or other related phenotypes. Finally, one point was awarded for tissue-specificity of mRNA expression reported in the Human Protein Atlas (41) (HPA) (proteinatlas.org) queried on 8/12/2024. If mRNA was enriched in reproductive tissues, brain, adipose, skeletal muscle, liver, or pancreas, this point was awarded.

**Figure 2.**
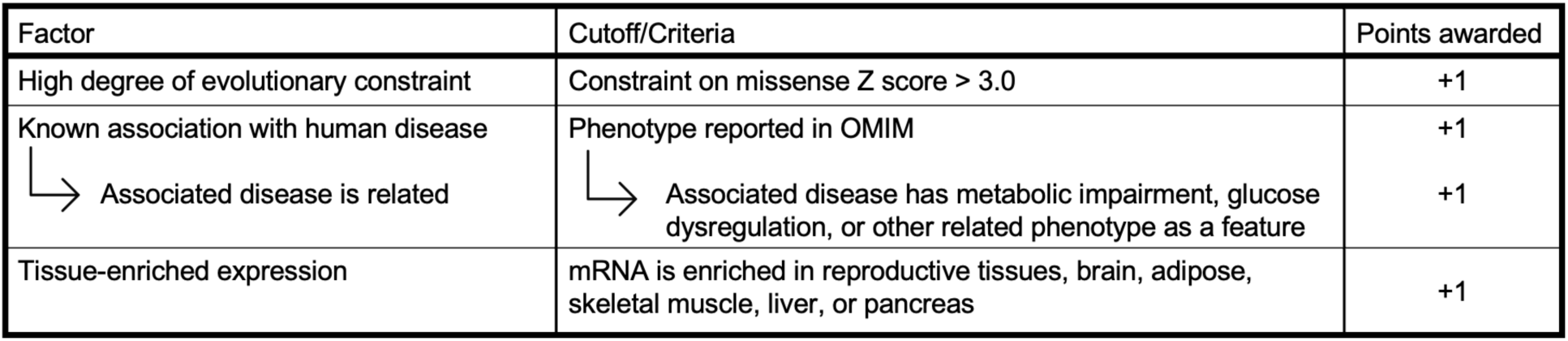
Candidate genes weighted confidence ranking strategy.

The 4-point system was used to create 5 categories of genes with varying degrees of confidence: 0 points as “none,” 1 point as “weak,” 2 points as “moderate,” 3 points as “strong,” and 4 points as “robust.” Genes with 2-4 points were selected for further investigation.

### Variant Filtering and Interpretation

Variants mapping to genes with at least moderate (2-4 points) evidence for candidate gene strength were filtered from the WES dataset containing protein-coding variants (missense, nonsense, frameshift). Variants with REVEL (38) score ≤ 0.6 and variants affecting control subjects only were eliminated from the dataset. In-frame insertions and deletions were assessed by the Sorting Intolerant From Tolerant (SIFT) coding indel classification algorithm (42), and variants considered “neutral” were discarded.

Variants selected for further analysis were classified using variant interpretation guidelines by the American College of Medical Genetics and Genomics (ACMG) and the Association for Molecular Pathology (AMP) (43). All missense variants in the dataset have REVEL ≥ 0.6 and therefore all have computational support for deleterious effect (ACMG/AMP-PP3). Nonsense and frameshift variants were predicted to be null (ACMG/AMP-PVS1). Genes with high constraint on missense scores (Z > 3.0) were expected to be genes with low rates of benign missense variants (ACMG/AMP-PP2). Statistical overrepresentation in PCOS cases compared to controls (ACMG/AMP-PS4) was assessed by OR using gnomAD v2.1.1 NFE population controls. An OR with a 95% confidence interval (CI) excluding 1 was considered significant. Functional data was assessed to determine if variants map to functional domains of the protein (ACMG/AMP-PM1) as defined in UniProt (uniprot.org) or had previous reports of deleterious effect in functional studies (ACMG/AMP-PS3). Previous reports of pathogenicity (ACMG/AMP-PS1) derived from literature search for case reports and/or genetic studies of disease cohorts were considered if related to the biology of IR.

### Selection of Highlighted Genes

Fifteen genes were selected for detailed variant analysis. To select the genes to highlight, genes were ranked on three scales: weight of evidence for candidacy, degree of association with PCOS, and number of PCOS cases affected by variants in the gene. A combined rank score from these three rankings was computed. The top 15 genes in the combined rank score were further investigated for their relationship to PCOS.

## Results

### Study participant characteristics

The PCOS cases in our study cohort had significantly higher testosterone levels (p<2×10^-16^) and BMI (p<2×10^-16^) than controls and did not differ significantly in age (Table 1).

**Table 1.** Clinical and biochemical features of PCOS cases and controls in this study.

|  | PCOS Cases (n=675) |  | Controls (n=235) |  | p value |
| --- | --- | --- | --- | --- | --- |
|  | n | Median (IQR) | n | Median (IQR) |  |
| Age (years) | 675 | 28 (25-32) | 235 | 26 (22-32) | n.s. |
| BMI (kg/m<sup>2</sup>) | 674 | 31.3 (24.8-39.0) | 234 | 23.1 (21.1-25.0) | <2x10<sup>-16</sup> |
| Testosterone (ng/dl) | 582 | 52 (37-72) | 224 | 32 (22-41) | <2x10<sup>-16</sup> |
| DHEAS (ng/ml) | 284 | 1950 (1313-2608) | 229 | 1930 (1375-2635) | n.s. |
| SHBG (nM) | 579 | 31.0 (20.5-46.0) | 228 | 61.0 (44.3-83.8) | 4.27x10<sup>-11</sup> |
| HOMA-IR | 593 | 2.02 (0.94-4.25) | 216 | 1.05 (0.71-1.44) | n.s. |
| WHR | 347 | 0.88 (0.83-0.93) | 270 | 0.82 (0.77-0.85) | <2x10<sup>-16</sup> |
| Fasting Glucose (mg/dl) | 612 | 87 (81-94) | 217 | 94 (87-101) | 1.91x10<sup>-7</sup> |
| Fasting Insulin (uIU/ml) | 609 | 9.3 (4.5-19.2) | 225 | 4.4 (3.1-6.3) | n.s. |
| HbA1c (%) | 306 | 5.3 (5.1-5.5) | 218 | 5.2 (5.0-5.4) | n.s. |
| Total cholesterol (mg/dl) | 635 | 179 (155-203) | 225 | 169 (150-192) | n.s. |
| HDL cholesterol (mg/dl) | 634 | 44 (35-56) | 225 | 61 (52-70) | 0.00754 |
| LDL cholesterol (mg/dl) | 633 | 113 (92-135) | 225 | 95 (80-111) | 3.97x10<sup>-4</sup> |
| Triglycerides (mg/dl) | 634 | 96 (66-143) | 225 | 59 (49-76) | 6.04x10<sup>-6</sup> |

Variables evaluated in a linear model assessing the contributions of age, BMI, case/control group, and assay or study site where applicable. P value represents the contribution of case/control group to the variation in the data.
Abbreviations: IQR, inter-quartile range; n.s., not significant; BMI, body-mass index; DHEAS, dehydroepiandrosterone sulfate; SHBG, sex hormone binding globulin; HOMA-IR, homeostatic model assessment of IR; WHR, waist-to-hip ratio; HbA1c, glycated hemoglobin A1c, HDL, high-density lipoprotein; LDL, low-density lipoprotein.

Metabolically, PCOS cases had lower SHBG (p=4.27×10^-11^) and HDL cholesterol (p=0.00754), and higher LDL cholesterol (p=3.97×10^-4^) and triglyceride levels (p=6.04×10^-6^) when compared to controls (Table 1). PCOS cases had higher waist-to-hip ratios (WHR) than controls (p<2×10^-16^). Fasting glucose levels were lower in PCOS cases than controls (p=5.04×10^-10^), which could indicate hyperinsulinemia preceding IR. PCOS cases had higher fasting insulin levels than controls, although the difference is not significant (Table 1). Overall, the phenotypic features of the PCOS cohort do not indicate an overtly insulin resistant subtype of PCOS; rather, they reflect a heterogenous, generic PCOS cohort.

### Gene Selection & Prioritization

Gene selection in the three IR gene sets yielded a total of 310 unique genes to be considered (Fig 1, Table S1). 235 genes comprised the insulin signaling gene set, 49 genes comprised the obesity gene set, and 26 genes comprised the dyslipidemia gene set. Two genes had duplicate rationale for inclusion within their gene set: *TBC1D4* was included in the insulin signaling gene set both by KEGG IR and T2D GWAS evidence, and *LEPR* was included in the obesity gene set both by monogenic obesity and obesity GWAS evidence (Table S1).

All 310 IR genes as well as the 100 randomly selected gene controls were assessed for their weighted strength as PCOS candidate genes (Table S2-S3). Just over half of the 100 randomly selected genes had no evidence for candidate weight, and only 16 of these genes received scores of moderate or above (Fig 3). In contrast, all three IR gene sets had higher proportions of genes receiving higher weighted confidence scores than the randomly selected genes (Fig 3). Sixteen of the randomly selected genes, 102 insulin signaling genes, 29 obesity genes, and 22 dyslipidemia genes received scores of moderate, strong, or robust. 153 IR genes across the three gene sets were therefore carried forward in the study (Fig 1, Table S2).

**Figure 3.**
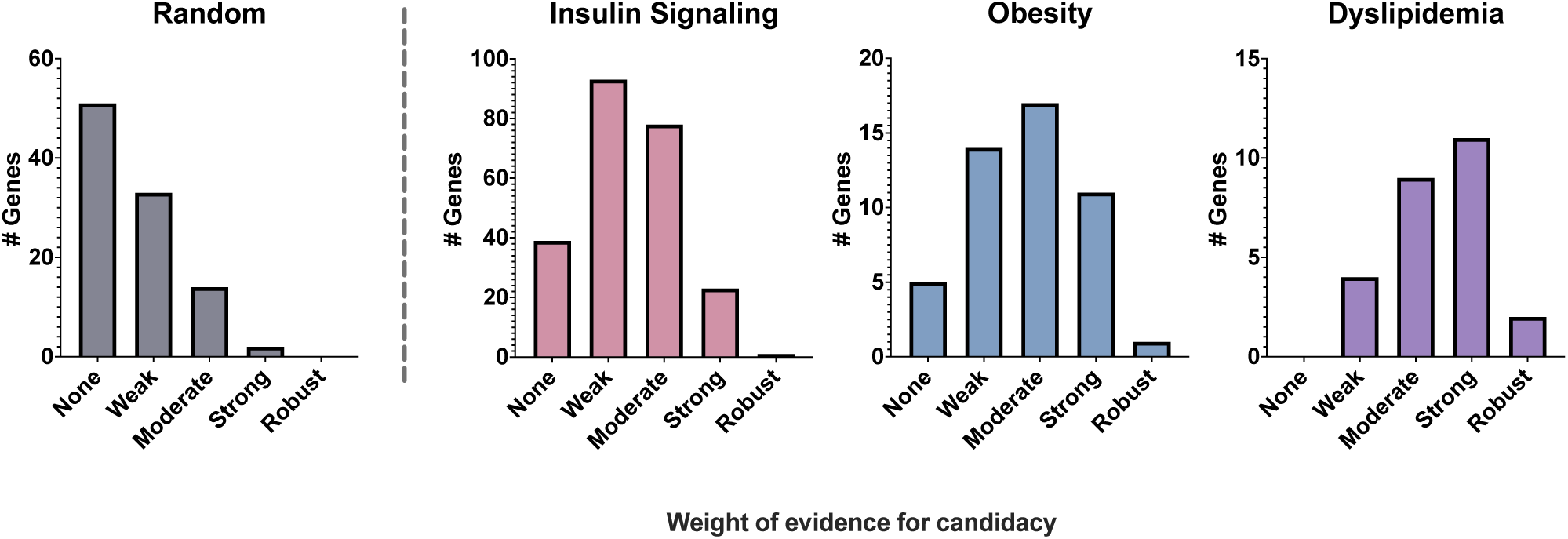
Distributions of weighted confidence rankings for each gene set. The number of genes with each level of weighted confidence was plotted to show the relative distribution of candidate strength. (None=0 points, Weak=1, Moderate=2, Strong=3, Robust=4)

### Prioritized Genes are Enriched for Variation in PCOS Cases

Nonsense, frameshift, and predicted damaging in-frame indels (SIFT indel “damaging”) and missense variants (REVEL ≥ 0.6) were identified in the 153 tested genes (Table S4). 59 genes did not contain any variants meeting these cutoffs. We identified 356 variants across 94 genes affecting 515 PCOS cases in our study (Fig 1, Table S4). We collapsed variants into association tests by gene to test for enrichment of variation in PCOS cases compared to population controls. 40 insulin signaling genes, 12 obesity genes, and 10 dyslipidemia genes were significantly enriched for variation in PCOS cases (Fig 1, 4, Table 2, Table S5).

**Table 2.**
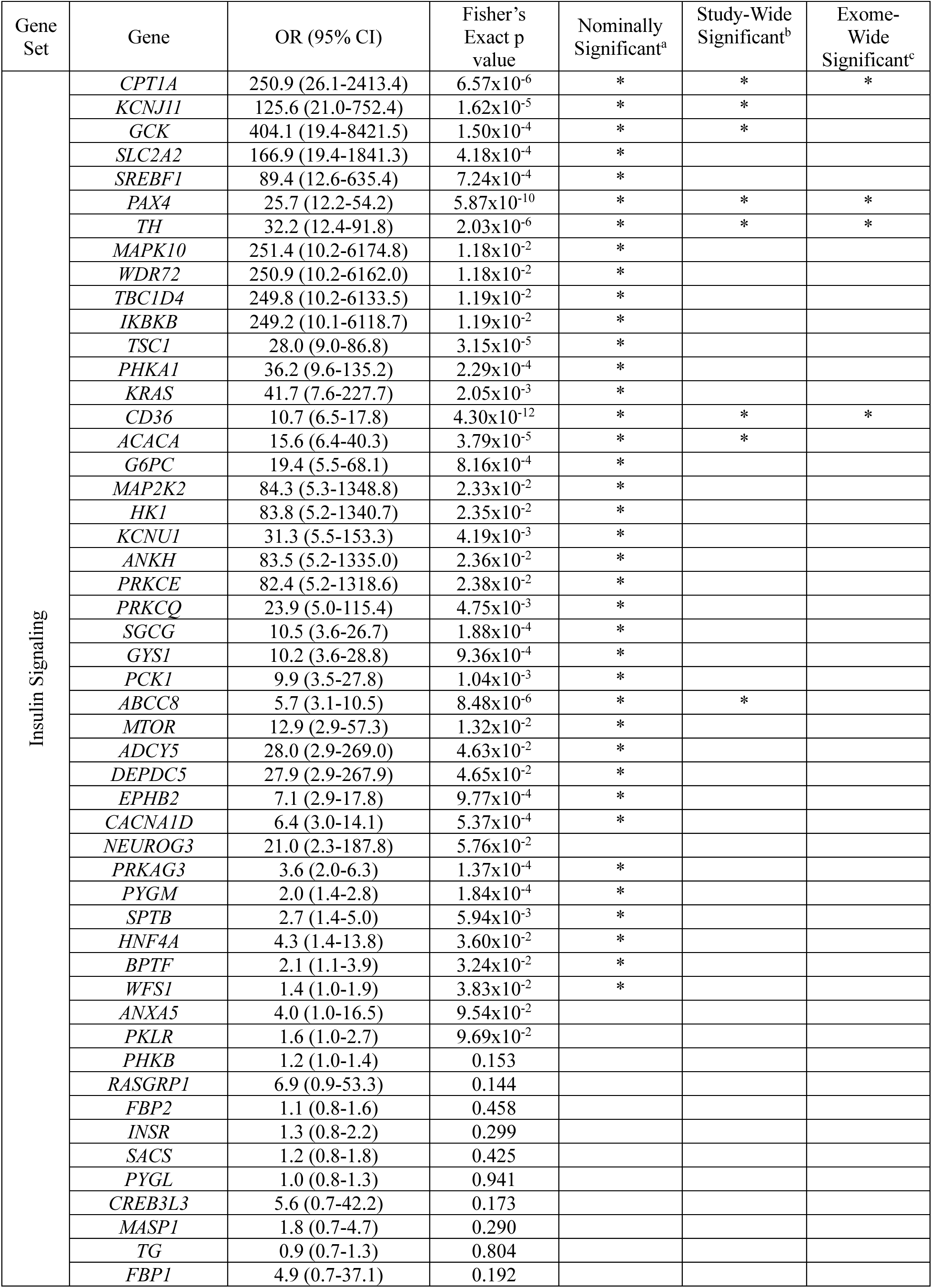
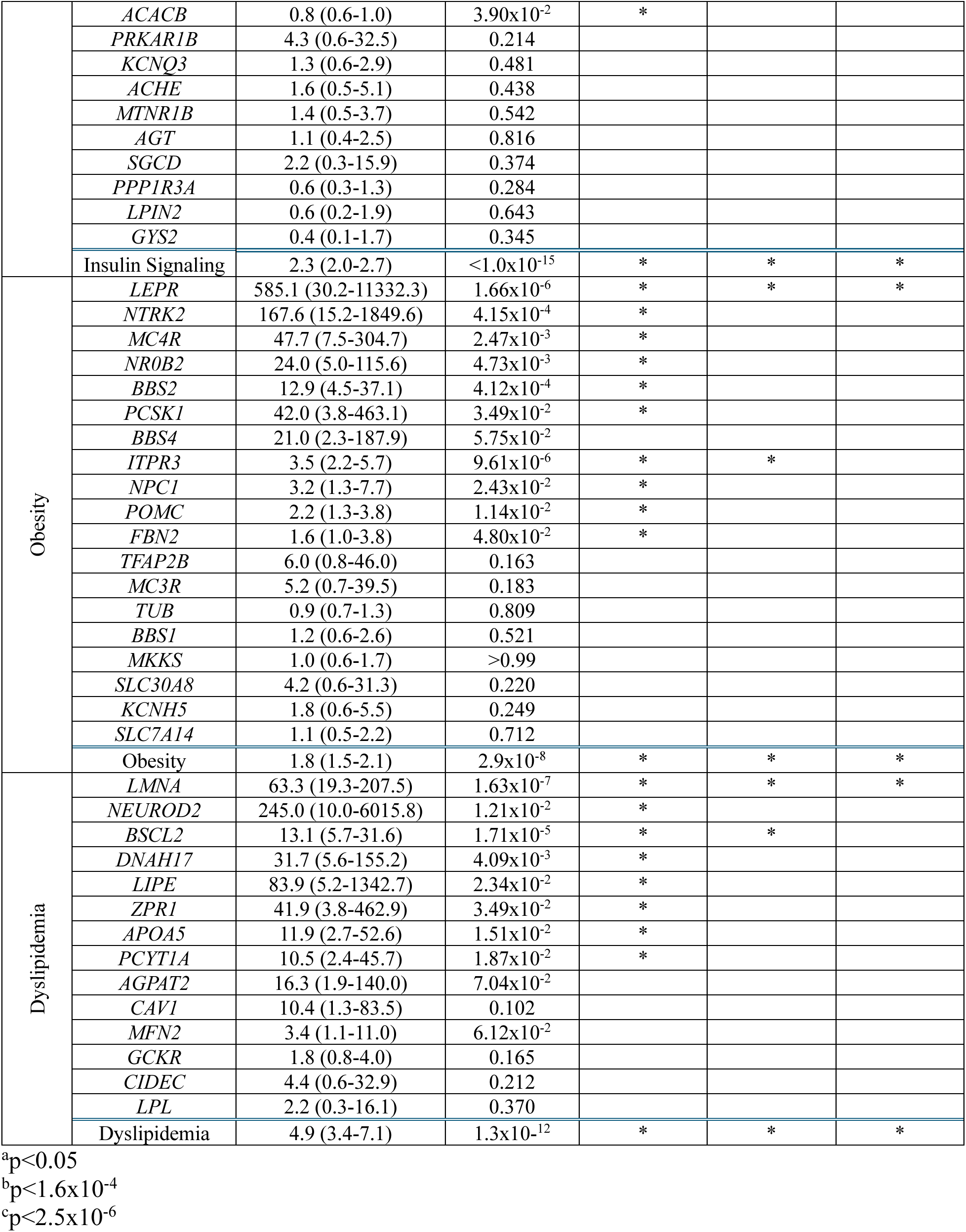
Gene-based associations for all genes assayed in this study.

**Table 3.**
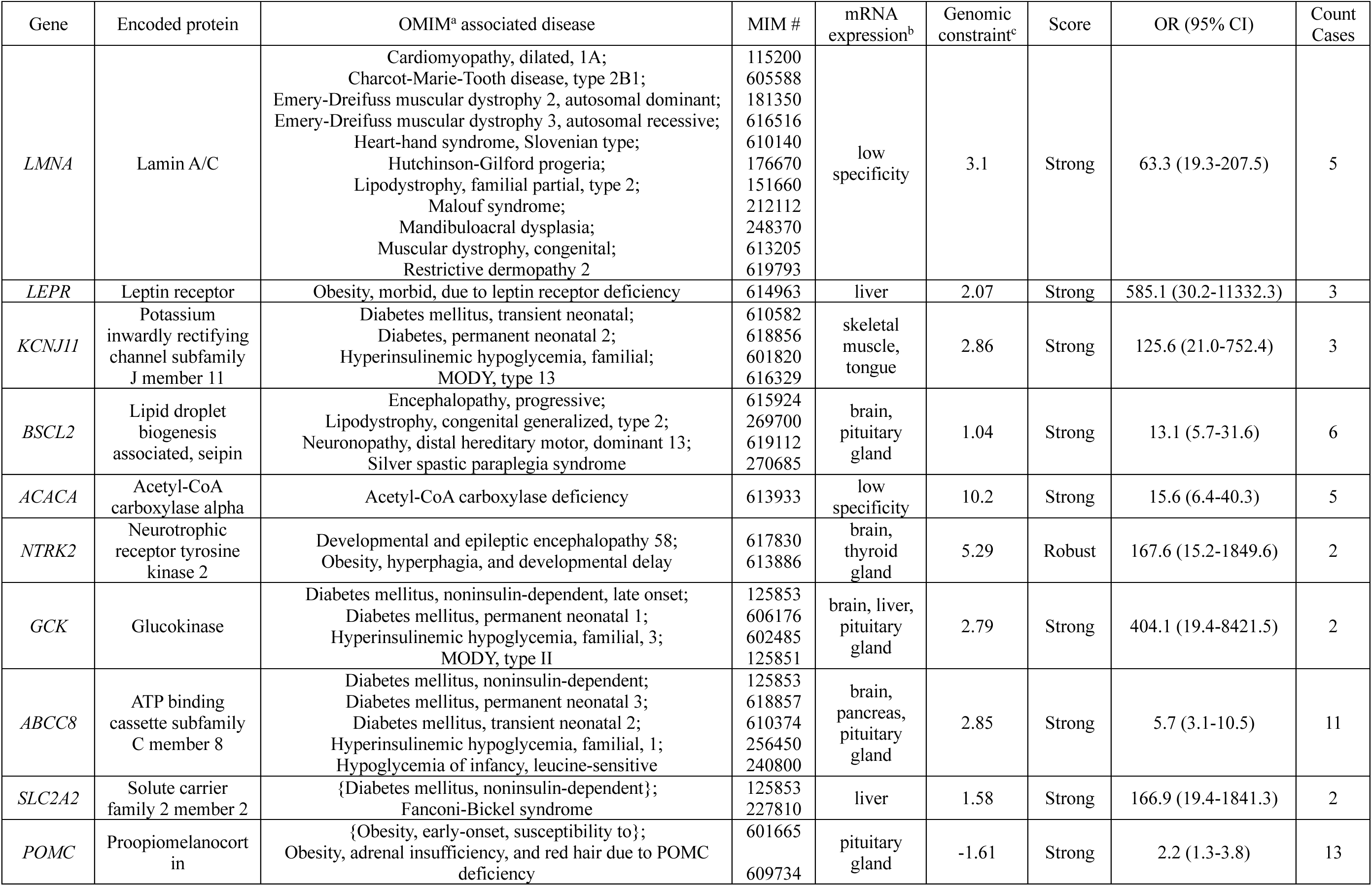
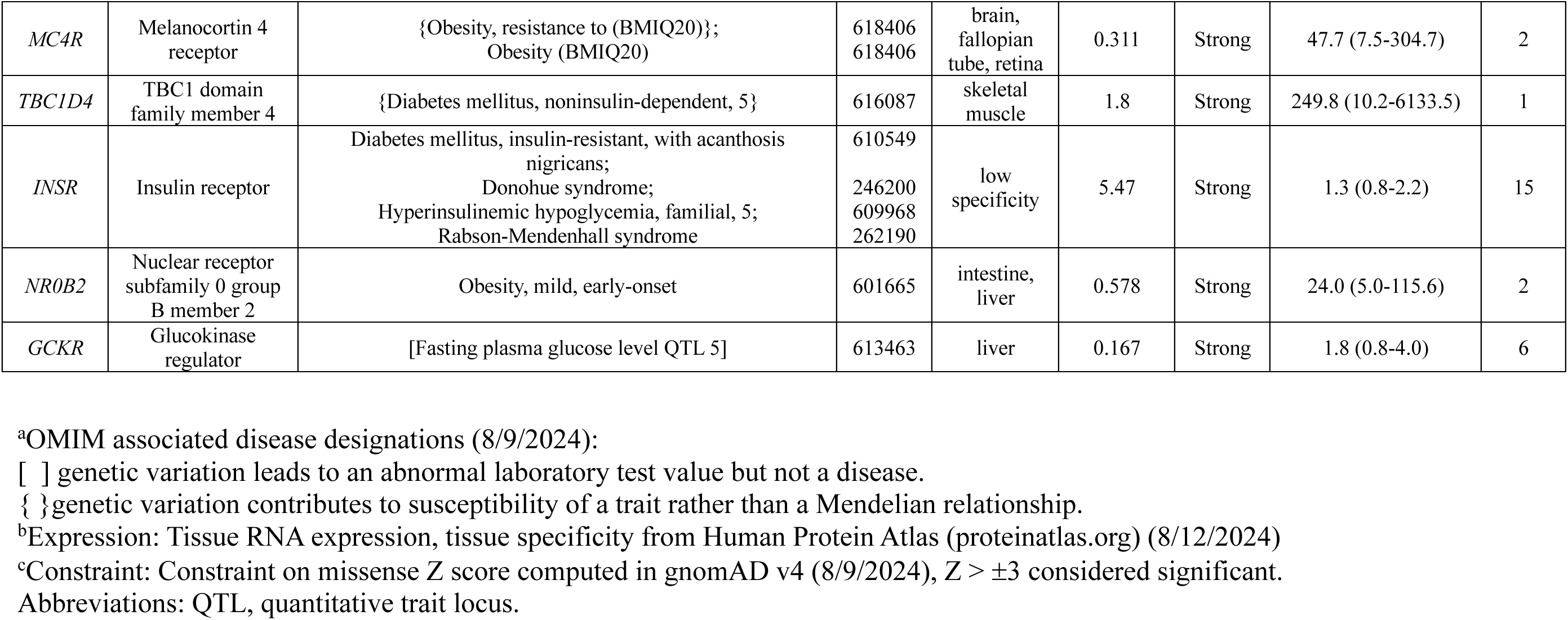
Candidate gene evidence and strength score for 15 highlighted PCOS genes.

Additionally, all three pathways are significantly enriched for variation in PCOS cases when variants were pooled by pathway, while the random genes were not (Fig 1, 4, Table 2). By Fisher’s exact test, 39 insulin signaling, 10 obesity, and 8 dyslipidemia genes were nominally significant (p<0.05) (Table 2). Four insulin signaling genes (*KCNJ11, GCK, ACACA, ABCC8*), one obesity gene (*ITPR3*), and one dyslipidemia gene (*BSCL2*) were significant at the study-wide level (p<1.6×10^-4^), and 4 insulin signaling genes (*CPT1A, PAX4, TH, CD36*), one obesity gene (*LEPR*), and one dyslipidemia gene (*LMNA*) were further significant at the exome-wide level (p<2.5×10^-6^) (Table 2, Fig 4). Variants in genes significantly associated with PCOS affected a total of 341 PCOS cases across all three IR gene sets, amounting to 51% of the PCOS cohort (Fig 1).

**Figure 4.**
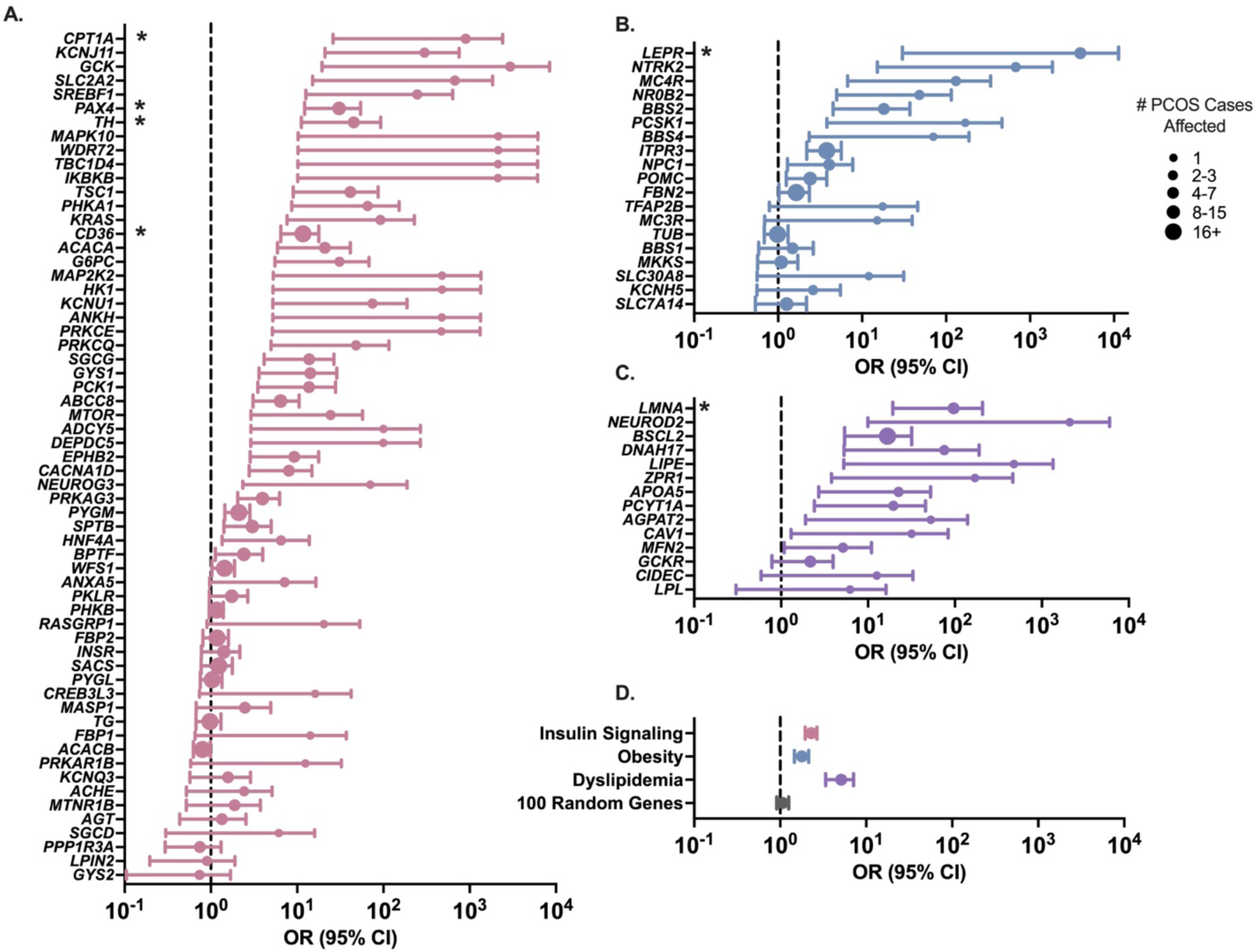
Enrichment of variation in PCOS cases compared to population controls. OR of PCOS cases relative to gnomAD non-Finnish European population controls and corresponding 95% CIs were plotted on a logarithmic axis. Genes are sorted by the lowest 95% CI limit and OR dots were sized according to the number of PCOS cases with a variant in the gene for gene-based ORs (A-C). (**A**) Insulin signaling, (**B**) Obesity, and (**C**) Dyslipidemia gene sets. Asterisks indicate genes significant at the exome-wide level (p<2.5×10^-6^) by Fisher’s exact test. (**D**) ORs computed as a pooled burden for each pathway.

### Top 15 genes selected to highlight

The 15 highest ranked genes were selected for in depth analysis. In these 15 genes, 48 variants were identified affecting 78 PCOS cases (Table 4-5, S6). 24 of these variants were classified as pathogenic, 17 as likely pathogenic, and only 7 as VUS (Table 5). Variants in the highlighted genes have been associated with a number of related pathologies, including dominant traits such as MODY (Leu135Val, Cys418Arg, Ala1536Val in *ABCC8*; Ala259Thr in *GCK*), and monogenic obesity (Thr150Ile in *MC4R*; His143Gln, Arg236Gly in *POMC*) and recessive traits such as congenital hyperinsulinemia (Gly92Ser, Leu135Val, Asp310Asn, Cys418Arg, Leu511Pro, Ala847Thr, and Ala1536Val in *ABCC8*; Ala96Val and Lys222Gln in *KCNJ11*) (Table 5).

**Table 4.**
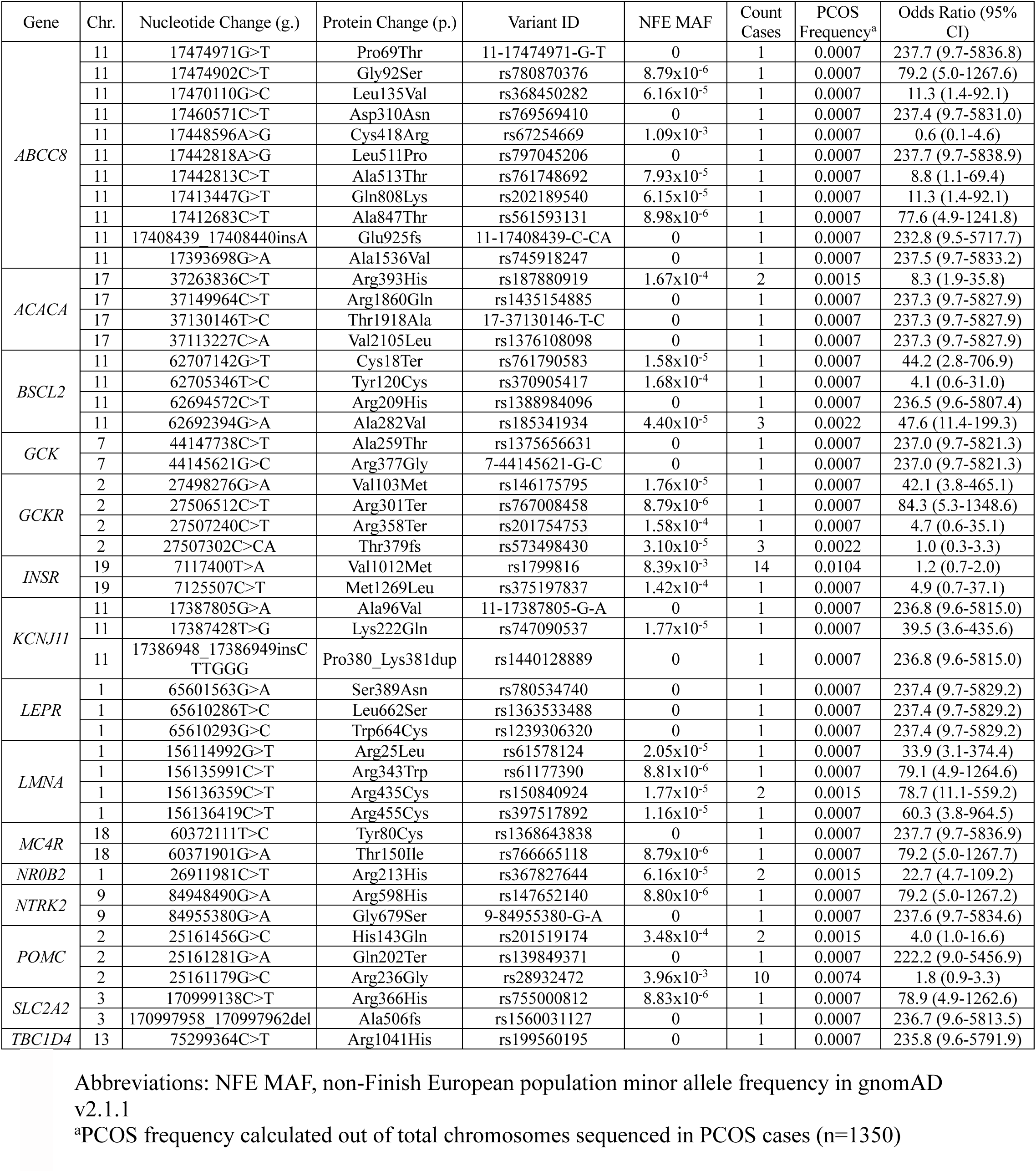
Variants identified in 15 highlighted PCOS genes.

**Table 5.** Effects of variants in 15 highlighted PCOS genes on protein function.

| Gene | Variant (p.) | REVEL Score | Affected functional protein domain | Previous Reports of Functional Impact | Previous Reports of Pathogenicity | Classification |
| --- | --- | --- | --- | --- | --- | --- |
| *ABCC8* | Pro69Thr | 0.891 |  |  | --- | VUS |
|  | Gly92Ser | 0.887 |  |  | CHI(84) | Pathogenic |
|  | Leu135Val | 0.689 |  | Impairs K<sub>ATP</sub> channel activity(85) | MODY,(86) CHI,(87) PAH(85) | Pathogenic |
|  | Asp310Asn | 0.833 | ABC transmembrane type-1 domain 1 | Decreases channel surface expression(88) | CHI(88-91) | Pathogenic |
|  | Cys418Arg | 0.657 | ABC transmembrane type-1 domain 1 |  | CHI,(92, 93) MODY,(94, 95) T2D(96) | Likely Pathogenic |
|  | Leu511Pro | 0.940 | ABC transmembrane type-1 domain 1 |  | CHI(97) | Pathogenic |
|  | Ala513Thr | 0.812 | ABC transmembrane type-1 domain 1 |  | T2D(98) | Pathogenic |
|  | Ile681Thr | 0.795 | ABC transporter domain 1 |  | --- | Likely Pathogenic |
|  | Gln808Lys | 0.602 | ABC transporter domain 1 |  | PAH,(99) T1D(100) | Pathogenic |
|  | Ala847Thr | 0.723 | ABC transporter domain 1 |  | CHI(101) | Pathogenic |
|  | Glu925fs | NA | ABC transporter domain 1 |  | --- | Likely Pathogenic |
|  | Ala1536Val | 0.951 | ABC transporter domain 2 |  | CHI,(102) MODY (Ala1536Thr)(103) | Pathogenic |
| *ACACA* | Arg393His | 0.888 | ATP-grasp domain |  | --- | Likely Pathogenic |
|  | Arg1860Gln | 0.908 | Carboxyl transferase domain |  | --- | Likely Pathogenic |
|  | Thr1918Ala | 0.601 | Carboxyl transferase domain |  | --- | Likely Pathogenic |
|  | Val2105Leu | 0.882 | Carboxyl transferase domain |  | --- | Likely Pathogenic |
| *BSCL2* | Cys18Ter | NA |  |  | --- | Likely Pathogenic |
|  | Tyr120Cys | 0.906 |  |  | --- | VUS |
|  | Arg209His | 0.837 |  |  | --- | VUS |
|  | Ala282Val | 0.784 |  |  | CMT(104) | Pathogenic |
| *GCK* | Ala259Thr | 0.901 | Hexokinase large subdomain |  | MODY,(105-111) T2D(112) | Pathogenic |
|  | Arg377Gly | 0.991 | Hexokinase large subdomain |  | MODY (Arg377His,(113, 114) Arg377Ser,(113) Arg377Leu,(115-117) Arg377Cys(118, 119)) | Likely Pathogenic |
| *GCKR* | Val103Met | 0.689 | Sugar isomerase (SIS) domain 1 | Severe loss of function in HeLa cells(120) | SHTG,(121-123) HLP1(124) | Pathogenic |
|  | Arg301Ter | NA |  |  |  | Likely Pathogenic |
|  | Arg358Ter | NA | Sugar isomerase (SIS) domain 2 |  |  | VUS |
|  | Thr379fs | NA | Sugar isomerase (SIS) domain 2 | Severe loss of function in HeLa cells(120) |  | Pathogenic |
| *INSR* | Val1012Met | 0.634 |  |  | T2D(125) | Likely Pathogenic |
|  | Met1269Leu | 0.830 | Protein tyrosine kinase domain |  | --- | VUS |
| *KCNJ11* | Ala96Val | 0.616 |  |  | CHI(98, 126) | Pathogenic |
|  | Lys222Gln | 0.714 |  |  | CHI(98) | Pathogenic |
|  | Pro380_Lys381dup | Damaging<sup>a</sup> |  |  | T2D(98) | Pathogenic |
| *LEPR* | Ser389Asn | 0.629 | Ig-like domain |  | Severe obesity,(127) HGTP(128) | Pathogenic |
|  | Leu662Ser | 0.732 | Fibronectin type-III domain 3 |  | Hyperphagia and severe obesity(129) | Pathogenic |
|  | Trp664Cys | 0.867 | Fibronectin type-III domain 3 |  | Hyperphagia and severe obesity (Trp664Arg)(130) | Likely Pathogenic |
| *LMNA* | Arg25Leu | 0.873 |  |  | FPLD2<sup>(131, 132)</sup> | Pathogenic |
|  | Arg343Trp | 0.683 | Intermediate filament rod domain |  | --- | Likely Pathogenic |
|  | Arg435Cys | 0.667 | Lamin tail domain |  | RD,<sup>(133, 134)</sup> HGPS<sup>(134, 135)</sup> | Pathogenic |
|  | Arg455Cys | 0.804 | Lamin tail domain |  | PCOS,<sup>(72)</sup> EDMD (R455P)<sup>(136)</sup> | Pathogenic |
| *MC4R* | Tyr80Cys | 0.722 |  |  | --- | VUS |
| | Thr150Ile | 0.685 | | Impairs receptor signaling in response to α-MSH binding(137-139) | Hyperphagia and severe obesity(137, 139-143) | Pathogenic |
| *NR0B2* | Arg213His | 0.887 |  |  | Mild obesity (Arg213Cys)(144) | VUS |
| *NTRK2* | Arg598His | 0.679 | Protein kinase domain |  | --- | Likely Pathogenic |
|  | Gly679Ser | 0.922 | Protein kinase domain |  | --- | Likely Pathogenic |
| *POMC* | His143Gln | 0.822 | α-MSH, corticotropin peptide region | | Severe early-onset obesity(145) | Pathogenic |
|  | Gln202Ter | NA |  |  | --- | Likely Pathogenic |
|  | Arg236Gly | 0.816 | Lipotropin beta, Met-enkephalin, Beta-endorphin peptide region | Prevents normal processing of POMC(146) | Severe early-onset obesity(145-151) | Pathogenic |
| *SLC2A2* | Arg366His | 0.843 |  |  | --- | Likely Pathogenic |
|  | Ala506fs | NA |  |  | --- | Likely Pathogenic |
| *TBC1D4* | Arg1041His | 0.602 | Rab-GAP TBC domain |  | Severe IR(152) | Pathogenic |

<sup>a</sup>Small insertion Pro380\_Lys381dup in *KCNJ11* was assessed with SIFT coding indel algorithm
rather than REVEL score.
Abbreviations: REVEL, rare exome variant ensemble learner; VUS, variant of uncertain significance;
CHI, congenital hyperinsulinemia; PAH, pulmonary arterial hypertension; T1D, type 1 diabetes; CMT,
Charcot-Marie-Tooth disease; SHTG, severe hypertriglyceridemia; HLP1, Type I
hyperlipoproteinemia; HGPT, hypertriglyceridemic pancreatitis; FPLD2, familial partial lipodystrophy
type 2; RD, restrictive dermopathy; HGPS, Hutchinson-Gilford progeria syndrome; EDMD, Emery-
Dreyfuss muscular dystrophy.

## Discussion

PCOS is a complex, multisystem genetic trait that is strongly associated with IR. To test the hypothesis that genetic forms of IR contribute to the etiology of PCOS, we evaluated genes in three categories of genetic IR for protein-altering variants in PCOS patients. From a list of 310 genes, we identified 153 genes with significant *a priori* evidence to warrant investigation. Out of these, 62 genes were significantly enriched for protein-altering variation in WES data from a cohort of 675 PCOS cases compared to population controls. Although individual variants are rare, 51% of PCOS cases were carriers of protein-altering variants in genes significantly enriched for variants compared to population controls, thus representing an incredibly common genetic feature of PCOS. These data provide support that for a significant subset of PCOS cases, IR may be a primary driver of the PCOS phenotype rather than a consequence of elevated testosterone levels.

To test the role of IR in PCOS in an unbiased and comprehensive manner but also maximize interpretability, we developed a gene curation and prioritization pipeline. The gene scoring system (Fig 2) was developed to optimize the identification of variants in genes amenable to clinical interpretation. We started with 310 genes in total and tested the genes with the strongest molecular and clinical evidence for a role in IR for further genetic evaluation. We evaluated 153 genes that survived our gene scoring system from the initial list of 310 genes, representing 49% of the genes initially considered. In contrast, out of 100 randomly selected genes scored alongside the IR gene sets, only 16 (16%) survived our prioritization screen. When we later examined the narrower set of 15 selected genes (Box 1) at the variant level, we did not identify any benign or likely benign variants, and only seven of 48 variants remained VUS (Table 5). By selecting genes with substantial evidence for disease association and high genomic constraint, and by using a protein functional prediction filter to select variants in those genes, our pipeline strongly enriched our results towards pathogenic variants. Our stringent gene prioritization and variant filtering system allowed us to clearly interpret the significance of variants in the final dataset. Although this may have resulted in a degree of type 2 error due to the exclusion of genes without strong of *a priori* evidence for a role in IR, this is preferable to type 1 error and the inclusion of false positive genes.

Both dominant and recessive forms of highly penetrant, monogenic diabetes, obesity, and dyslipidemia have been identified previously, but it was unclear whether the same genes or causal variants are also implicated in PCOS, a common, complex trait. Our gene curation strategy included genes with OMIM associations of MODY, monogenic obesity, lipodystrophy, or other syndromic pathologies with obesity or IR as a feature. We assayed 41 genes related to known monogenic manifestations of IR from the WES data collected for our PCOS cohort. Of these, 29 genes contained at least one variant surviving our functional filter in least one PCOS case and 21 of these genes showed significant association with PCOS. These include the MODY genes *KCNJ11, GCK, HNF4A* and *PAX4*, familial hyperinsulinemic hypoglycemia gene *ABCC8*, monogenic obesity genes *LEPR, NR0B2, POMC, MC4R,* and *NTRK2,* Bardet-Biedl syndrome genes *BBS2* and *BBS4*, partial lipodystrophy genes *LMNA, LIPE* and *MFN2*, and generalized lipodystrophy genes *BSCL2, AGPAT2*, *CAV1*, and *PCYT1A*. Along with our previous study of PCOS patients with pathogenic variants in *LMNA* (16), this further underscores that within the heterogeneous group of individuals diagnosed with PCOS, some may have distinct monogenic etiologies. This is essential to understanding the pathogenesis of PCOS, especially in the context of genetic causes of obesity and IR. Accurate and effective treatment of PCOS depends on recognizing obesity and IR as potential primary drivers of the condition and thus targets for treatment.

Our results add to a growing body of studies that challenge classical inheritance models especially in the context of common, complex traits (44). It is possible that the features of PCOS include symptomatic heterozygous forms of recessive traits like generalized lipodystrophy or severe IR. If variants can only exert their effects in the recessive state, we would expect their prevalence in population databases to be too common to result in statistical enrichment in our cohort. In contrast, numerous genes significantly enriched for variation in PCOS cases in our study are in fact related to recessive disorders of IR. Examples include *ABCC8* (familial hyperinsulinemic hypoglycemia), *BSCL2, PCYT1A, CAV1,* and *AGPAT2* (generalized lipodystrophy), *BBS2* and *BBS4* (Bardet-Biedl syndrome), *SLC2A2* (Fanconi-Bickel syndrome), *ACACA* (Acetyl-CoA carboxylase deficiency), *NPC1* (Niemann-Pick disease), and *G6PC* and *GYS1* (glycogen storage disorders) (Table S2). The prevalence of heterozygous protein-altering variants in these genes in our PCOS cohort supports possible additive inheritance for these genes, or an otherwise symptomatic heterozygote model.

Although the insulin signaling, obesity, and dyslipidemia gene sets were curated and stratified using the same strategy, there were clear differences in how each gene set contributes to the heritability of PCOS. The obesity and dyslipidemia sets were primarily defined by genes already demonstrated to be associated with disease, while the insulin signaling gene set was largely selected by KEGG molecular pathway designations. We expected that our prioritization pipeline would differentially weight genes in the disease-defined gene sets higher; in fact, 57% of insulin signaling genes were eliminated before assaying from the sequencing dataset, whereas only 41% of obesity genes and 15% of dyslipidemia genes were eliminated (Fig 1). This was also reflected in the distributions of genes receiving each level of supporting evidence (Fig 3), where the insulin signaling gene set has a right-skewed distribution (most genes receiving low scores), the obesity gene set is nearly normally distributed, and the dyslipidemia gene set has a left-skewed distribution of evidence (most genes receiving high scores). In contrast, however, 84% of the 100 randomly selected genes were eliminated before assaying from the sequencing dataset, and the distribution of genes receiving each level of supporting evidence was extremely skewed towards genes receiving low scores (Fig 3). This underscores that while the insulin signaling gene set appears the weakest out of the IR gene sets, it is still starkly different from unselected, arbitrary genes. It is possible that variants in genes in the insulin signaling gene set have a smaller effect size than variants in dyslipidemia genes, with variants in obesity genes falling in the middle. The full effects of these variants may require polygenic inheritance or epistatic interactions within the pathway.

While our unbiased gene curation and filtering strategy was designed to reduce noise and increase interpretability, it may have introduced biases towards highly studied genes and rare variants. To mitigate the sequence variant interpretation bottleneck, we implemented rigorous pre-processing of genes, largely informed by the pieces of evidence that are incorporated in the ACMG variant curation (43). These criteria heavily favor genes associated with rare, monogenic disease and may not encompass genes contributing to PCOS heritability through more common, polygenic inheritance. Additionally, at the variant level, we applied a relatively stringent functional filter (REVEL ≥ 0.6). We chose this approach over a population frequency filter for two reasons: 1) to limit the inclusion of *de novo* variants that while stochastically rare, are not necessarily selected against and may not have clear functional impacts; and 2) to prevent the possible erroneous exclusion of relatively common variants with predicted functional effects.

Our approach did, however, still yield almost entirely rare variation; the highest NFE MAF of a single variant in the REVEL-filtered dataset was 0.0633. These biases towards well-known genes and rare variants served the important purpose of increasing interpretability, but future efforts will need to expand and begin identifying the “medium-hanging fruit” type of variation that contributes to pathogenesis in ways other than rare, high-effect causes. These types of variants may require molecular evaluation for their effects to be clear.

We have demonstrated that genetic causes of IR may contribute to the heritability of PCOS in a large proportion of women, and that a candidate pathway-driven approach to focus WES data can yield manageable, interpretable data. The selected gene sets of insulin signaling, obesity, and dyslipidemia may contribute to PCOS etiology through distinct mechanisms, but all represent important areas of further investigation into disease pathogenesis. For example, the genes identified as significantly enriched for variation in PCOS cases in this study and the molecular pathways they affect are prime candidates for functional screens or mechanistic studies to clarify their causal roles. These genes and pathways may merit follow-up through whole-genome sequencing to investigate variation in regulatory regions or deep intronic sites not captured by WES. Noncoding variation has been proposed as a likely causal mechanism for common, complex traits like PCOS, but whole-genome sequencing analysis requires even greater focusing of the data to reach meaningful results. Our gene discovery strategy may be useful to identify novel genes that can subsequently be investigated in greater detail, such as for regulatory region or splice site variants. More broadly, our approach can be applied to other functional pathways that may underlie PCOS pathogenesis and other complex genetic traits. Ultimately, this work advances our understanding of the complex etiology of PCOS and continues to move the field towards personalized medicine and treatments targeted at the precise underlying causes of disease.

## Data Availability

All data produced in the present study are available upon reasonable request to the authors

## Acknowledgements

We thank all individuals who participated in this study either as PCOS cases or healthy controls. Figure 1 was created with BioRender.com. This study was supported by US National Institutes of Health (NIH) grants R01 HD057450 (MU), P50 HD044405 (MU), R01 HD100630 (MU), R01 HD056510 (RSL), R01 HD065029 (CKW) and T32 HD094699 (RB). Additional funding was provided by the Androgen Excess & PCOS Foundation Waterloo Award (RB). Partial funding for the clinical studies was provided by UL1 TR000150, UL1 RR033184, UL1 TR000430 and UL1 RR025758 from the National Center for Advancing Translational Sciences. Some hormone assays were performed at the University of Virginia Center for Research in Reproduction Ligand Assay and Analysis Core that is supported by U54 HD28934 from the Eunice Kennedy Shriver National Institute of Child Health and Human Development.

## Author Contributions

Conceptualization: RB, ZCM, MGH, CKW, RSL, MU. Data curation: RB, CP, MGH, ARK. Formal analysis: RB. Resources: CKW, RSL. Funding Acquisition: MU, CKW, RSL, RB. Supervision: MGH, MU. Writing – Original Draft: RB, MU. Writing – Review & Editing: RB, CP, ZCM, MGH, ARK, RSL, CKW, MU.

## Box 1

***ABCC8*** encodes the SUR1 protein, part of an ATP-sensitive potassium channel that regulates insulin release from the pancreas. It has been implicated in T2D through GWAS, and rare variants also cause familial hyperinsulinemic hypoglycemia. Common variants associated with T2D have been shown to lack association with PCOS (45), but the coding region has not been assessed in the PCOS context.

***ACACA*** encodes a member of the insulin signaling pathway that regulates de novo fatty acid synthesis. Homozygous variants in this gene cause a rare metabolic syndrome called Acetyl-CoA carboxylase deficiency. It has not previously been a candidate gene for PCOS, but has known roles in oocyte maturation (46) and ovarian steroidogenesis (47).

***BSCL2***, also known as seipin, encodes an essential regulator of lipid droplet formation and adipocyte development. Variants in this gene cause CGL2, a recessive lipodystrophy syndrome. A relationship between CGL2 and PCOS was first proposed nearly 50 years ago (48), but only sporadic case reports document symptoms of PCOS in CGL2 patients,(49) and a previous sequencing effort in a PCOS cohort did not identify variants in *BSCL2* (15).

***GCK*** encodes the glucokinase enzyme, a member of the insulin signaling network whose dysfunction causes MODY and other forms of diabetes mellitus. Although at least one group has included *GCK* on a candidate sequencing panel (15), no variants have been associated with PCOS before.

***GCKR*** encodes glucokinase regulatory protein. It qualified for inclusion in this study due to a GWAS association with triglyceride level but also has an OMIM association with fasting plasma glucose levels, suggesting a multifaceted role in glucose homeostasis and IR. One previous study examined common copy-number variation in *GCKR* in PCOS cases with little success (50), and no studies have sequenced *GCKR* for coding variants in PCOS until now.

***INSR***, which encodes the insulin receptor, has long been a candidate gene for PCOS (8, 51–62). However, its candidacy has been based on associations with common, noncoding variants. Since its most penetrant related diseases Donohue syndrome and Rabson-Mendenhall syndrome are recessive disorders, *INSR* may not be an ideal candidate gene for rare causal variants in the heterozygous state.

***KCNJ11*** encodes an ATP-sensitive potassium channel essential for regulating insulin release from pancreatic beta cells. It has long been a GWAS candidate for T2D, and rare variants in this gene cause familial hyperinsulinemic hypoglycemia and MODY. Multiple studies have tested for association between common T2D polymorphisms and PCOS and found no relationship (25, 26, 63), but rare variation has not been investigated until the present work.

***LEPR*** encodes the leptin receptor, a key regulator of appetite and fat metabolism expressed in the liver. Common variants in *LEPR* have been associated with BMI in GWAS, and rare variants have been shown to cause monogenic morbid obesity. Common variants have been associated with PCOS in several populations (27, 28, 64–69), but a previous attempt to sequence the gene for rare variation in a PCOS cohort did not yield any variants (15).

***LMNA*** encodes the lamin A/C intermediate filament proteins expressed in all differentiated cell types. Because of variants in this gene that cause FPLD2, a syndrome with overlapping features of hyperandrogenism and menstrual dysfunction, *LMNA* has been a prime candidate gene for PCOS. Several studies have identified rare pathogenic variants in *LMNA* in women with PCOS (15, 16, 20, 70–72), cementing it as a reproducible candidate gene (73).

***MC4R***, which encodes a melanocortin receptor primarily expressed in the brain, is a preeminent obesity gene due to GWAS associations and high effect, monogenic cases alike. Common *MC4R* variants have been tested for association with PCOS in multiple studies, but have found no associations with PCOS itself, only with BMI/obesity (74–77).

***NR0B2***, also known as SHP, encodes an orphan receptor expressed in the liver and GI tract with proposed roles in adipogenesis and cholesterol metabolism. Variants in this gene cause mild early onset obesity and have never been reported in the context of PCOS. In one study of gene expression in adipocytes differentiated from human embryonic stem cells derived from PCOS patients and controls, *NR0B2* expression was found to be higher in PCOS-derived adipocytes than those from controls (78).

***NTRK2***, also named *TRKB,* encodes a tyrosine kinase receptor (TrkB) that binds its ligand brain-derived neurotrophic factor (BDNF). It is expressed in the brain and variants are associated with hyperphagic obesity. Although it hasn’t been considered a genetic target in PCOS before, several functional studies have established a role for BDNF-TrkB signaling in germ cell nest breakdown (79), oocyte maturation (80), and follicle development (81).

***POMC*** encodes a precursor peptide expressed in the pituitary that gets processed into several peptide hormones, including the melanocortins. Its dysfunction causes hyperphagic obesity and has not yet been shown to have a direct role in PCOS, although one study did include it in their gene panel but did not identify any variants (15).

***SLC2A2*** encodes the GLUT-2 glucose transporter expressed primarily in the liver. Variants in this gene cause Fanconi-Bickel syndrome, a glycogen storage disease, as well as susceptibility to T2D. Two studies have considered the role of *SLC2A2* in PCOS: one found a strong association between PCOS and a common variant (29), and the other sought to associate noncoding SNPs with metformin response (82).

***TBC1D4***, also known as AS160, encodes a substrate of AKT phosphorylation downstream of insulin receptor activation. Common variation in *TBC1D4* is associated with T2D. One study has shown a lesser presence of AS160 phosphorylation in muscle biopsies of PCOS patients compared to controls (83), but no studies have sequenced *TBC1D4* in PCOS cases until now.

## References

1. Legro RS, et al. Diagnosis and Treatment of Polycystic Ovary Syndrome: An Endocrine Society Clinical Practice Guideline. J Clin Endocrinol Metab. 2013;98(12):4565–4592.

2. Group REASPCW. Revised 2003 consensus on diagnostic criteria and long-term health risks related to polycystic ovary syndrome (PCOS). Hum Reprod. 2004;19(1):41-47.

3. Azziz R, et al. The Androgen Excess and PCOS Society criteria for the polycystic ovary syndrome: the complete task force report. Fertil Steril. 2009;91(2):456–488.

4. Diamanti-Kandarakis E, and Dunaif A. Insulin resistance and the polycystic ovary syndrome revisited: an update on mechanisms and implications. Endocr Rev. 2012;33(6):981–1030.

5. Vink J, et al. Heritability of polycystic ovary syndrome in a Dutch twin-family study. J Clin Endocrinol Metab. 2006;91(6):2100–2104.

6. Jahanfar S, et al. A twin study of polycystic ovary syndrome. Fertil Steril. 1995;63(3):478–486.

7. Hayes MG, et al. Genome-wide association of polycystic ovary syndrome implicates alterations in gonadotropin secretion in European ancestry populations. Nat Comm. 2015;6(1):7502.

8. Tian Y, et al. PCOS-GWAS Susceptibility Variants in THADA, INSR, TOX3, and DENND1A Are Associated With Metabolic Syndrome or Insulin Resistance in Women With PCOS. Front Endocrinol (Lausanne). 2020;11.

9. Manolio TA, et al. Finding the missing heritability of complex diseases. Nature. 2009;461(7265):747-753.

10. Andrews SJ, et al. The complex genetic architecture of Alzheimer’s disease: novel insights and future directions. eBioMedicine. 2023;90.

11. Cho MH, et al. Genetics of chronic obstructive pulmonary disease: understanding the pathobiology and heterogeneity of a complex disorder. Lancet Respir Med. 2022;10(5):485–496.

12. Lalami I, et al. Genomics of Endometriosis: From Genome Wide Association Studies to Exome Sequencing. Int J Mol Sci. 2021;22(14):7297.

13. Defesche JC, et al. Familial hypercholesterolaemia. Nat Rev Dis Primers. 2017;3(1):1–20.

14. Nkonge KM, et al. The epidemiology, molecular pathogenesis, diagnosis, and treatment of maturity-onset diabetes of the young (MODY). Clin Diabetes Endocrinol. 2020;6:1–10.

15. Crespo RP, et al. High-throughput Sequencing to Identify Monogenic Etiologies in a Preselected Polycystic Ovary Syndrome Cohort. J Endocr Soc. 2022;6(9).

16. Bauer R, et al. Rare Variation in LMNA Underlies Polycystic Ovary Syndrome Pathogenesis in 2 Independent Cohorts. J Clin Endocrinol Metab. 2024:dgae761.

17. Gorsic LK, et al. Pathogenic Anti-Mullerian Hormone Variants in Polycystic Ovary Syndrome. J Clin Endocrinol Metab. 2017.

18. Gorsic LK, et al. Functional genetic variation in the anti-Müllerian hormone pathway in women with polycystic ovary syndrome. J Clin Endocrinol Metab. 2019;104(7):2855–2874.

19. Hegele RA. Monogenic forms of insulin resistance: apertures that expose the common metabolic syndrome. Trends Endocrinol Metab. 2003;14(8):371–377.

20. Vantyghem MC, et al. Fertility and Obstetrical Complications in Women with LMNA-Related Familial Partial Lipodystrophy. J Clin Endocrinol Metab. 2008;93(6):2223–2229.

21. Marian AJ. The Bottleneck in Genetic Testing. Circ Res. 2015;117(7):586–588.

22. Burke W, et al. The Challenge of Genetic Variants of Uncertain Clinical Significance: A Narrative Review. Ann Intern Med. 2022;175(7):994–1000.

23. Petersen MC, and Shulman GI. Mechanisms of Insulin Action and Insulin Resistance. Physiol Rev. 2018;98(4):2133–2223.

24. Semple RK, et al. Genetic syndromes of severe insulin resistance. Endocr Rev. 2011;32(4):498–514.

25. Barber TM, et al. Relationship between E23K (an established type II diabetes-susceptibility variant within KCNJ11), polycystic ovary syndrome and androgen levels. Eur J Hum Genet. 2007;15(6):679–684.

26. Kim JJ, et al. Polycystic ovary syndrome is not associated with polymorphisms of the TCF7L2, CDKAL1, HHEX, KCNJ11, FTO and SLC30A8 genes. Clin Endocrinol (Oxf). 2012;77(3):439-445.

27. Li L, et al. Relationship between leptin receptor and polycystic ovary syndrome. Gene. 2013;527(1):71–74.

28. Wang N-N, et al. Association between leptin receptor polymorphisms and polycystic ovary syndrome risk: a meta-analysis based on 11 studies. Gynecol Endocrinol. 2023;39(1):2279565.

29. Alsobaie S, et al. Examining the Genetic Role of rs8192675 Variant in Saudi Women Diagnosed with Polycystic Ovary Syndrome. Diagnostics. 2023;13(20):3214.

30. Welt CK, et al. Defining Constant Versus Variable Phenotypic Features of Women with Polycystic Ovary Syndrome Using Different Ethnic Groups and Populations. J Clin Endocrinol Metab. 2006;91(11):4361–4368.

31. Welt C, et al. Characterizing discrete subsets of polycystic ovary syndrome as defined by the Rotterdam criteria: the impact of weight on phenotype and metabolic features. J Clin Endocrinol Metab. 2006;91(12):4842–4848.

32. Legro RS, et al. The Pregnancy in Polycystic Ovary Syndrome II (PPCOS II) trial: rationale and design of a double-blind randomized trial of clomiphene citrate and letrozole for the treatment of infertility in women with polycystic ovary syndrome. Contemp Clin Trials. 2012;33(3):470–481.

33. Legro RS, et al. The Pregnancy in Polycystic Ovary Syndrome II study: baseline characteristics and effects of obesity from a multicenter randomized clinical trial. Fertil Steril. 2014;101(1):258–269.e258.

34. ESHRE TR, and Group A-SPCW. Revised 2003 consensus on diagnostic criteria and long-term health risks related to polycystic ovary syndrome. Fertil Steril. 2004;81(1):19-25.

35. Cingolani P, et al. A program for annotating and predicting the effects of single nucleotide polymorphisms, SnpEff: SNPs in the genome of Drosophila melanogaster strain w1118; iso-2; iso-3. fly. 2012;6(2):80-92.

36. Cingolani P, et al. Using Drosophila melanogaster as a model for genotoxic chemical mutational studies with a new program, SnpSift. Front Genet. 2012;3:35.

37. Karczewski KJ, et al. The mutational constraint spectrum quantified from variation in 141,456 humans. Nature. 2020;581(7809):434-443.

38. Ioannidis NM, et al. REVEL: An Ensemble Method for Predicting the Pathogenicity of Rare Missense Variants. Am J Hum Genet. 2016;99(4):877–885.

39. Amberger JS, et al. OMIM.org: Online Mendelian Inheritance in Man (OMIM®), an online catalog of human genes and genetic disorders. Nucleic Acids Res. 2014;43(D1):D789–D798.

40. Chen S, et al. A genomic mutational constraint map using variation in 76,156 human genomes. Nature. 2024;625(7993):92-100.

41. Uhlén M, et al. Tissue-based map of the human proteome. Science. 2015;347(6220):1260419.

42. Ng PC, and Henikoff S. Predicting deleterious amino acid substitutions. Genome Res. 2001;11(5):863–874.

43. Richards S, et al. Standards and guidelines for the interpretation of sequence variants: a joint consensus recommendation of the American College of Medical Genetics and Genomics and the Association for Molecular Pathology. Genet Med. 2015;17(5):405–424.

44. Kalyta K, et al. The Spectrum of the Heterozygous Effect in Biallelic Mendelian Diseases—The Symptomatic Heterozygote Issue. Genes. 2023;14(8):1562.

45. Barber T, et al. Association between polymorphisms within the SUR1/Kir6. 2 gene region and polycystic ovary syndrome: a case-control study. Endocr Abstr. 2006;11.

46. Valsangkar DS, and Downs SM. Acetyl CoA carboxylase inactivation and meiotic maturation in mouse oocytes. Mol Reprod Dev. 2015;82(9):679–693.

47. Tosca L, et al. Effects of Metformin on Bovine Granulosa Cells Steroidogenesis: Possible Involvement of Adenosine 5′ Monophosphate-Activated Protein Kinase (AMPK)1. Biol Reprod. 2007;76(3):368–378.

48. Huseman CA, et al. Congenital lipodystrophy. II. Association with polycysticovarian disease. J Pediatr. 1979;95(1):72–74.

49. Shirwalkar HU, et al. Congenital generalized lipodystrophy in an Indian patient with a novel mutation in BSCL2 gene. J Inherit Metab Dis. 2008;31(2):317–322.

50. Knebel B, et al. Association between copy-number variation on metabolic phenotypes and HDL-C levels in patients with polycystic ovary syndrome. Mol Biol Rep. 2017;44(1):51–61.

51. Lee E-J, et al. Single nucleotide polymorphism in exon 17 of the insulin receptor gene is not associated with polycystic ovary syndrome in a Korean population. Fertil Steril. 2006;86(2):380–384.

52. Lee E-J, et al. A novel single nucleotide polymorphism of INSR gene for polycystic ovary syndrome. Fertil Steril. 2008;89(5):1213–1220.

53. Feng C, et al. The Association between Polymorphism of INSR and Polycystic Ovary Syndrome: A Meta-Analysis. Int J Mol Sci. 2015;16(2):2403–2425.

54. M TM, et al. INSR gene variation is associated with decreased insulin sensitivity in Iraqi women with PCOs. Iran J Reprod Med. 2014;12(7):499–506.

55. Seyed Abutorabi E, et al. Investigation of the FSHR, CYP11, and INSR Mutations and Polymorphisms in Iranian Infertile Women with Polycystic Ovary Syndrome (PCOS). Rep Biochem Mol Biol. 2021;9(4):470-477.

56. Thangavelu M, et al. Single-nucleotide polymorphism of INS, INSR, IRS1, IRS2, PPAR-G and CAPN10 genes in the pathogenesis of polycystic ovary syndrome. J Genet. 2017;96(1):87-96.

57. Siegel S, et al. A C/T single nucleotide polymorphism at the tyrosine kinase domain of the insulin receptor gene is associated with polycystic ovary syndrome. Fertil Steril. 2002;78(6):1240–1243.

58. Chen Z-j, et al. [Correlation between single nucleotide polymorphism of insulin receptor gene with polycystic ovary syndrome]. Zhonghua fu chan ke za zhi. 2004;39(9):582–585.

59. Mukherjee S, et al. Genetic variation in exon 17 of INSR is associated with insulin resistance and hyperandrogenemia among lean Indian women with polycystic ovary syndrome. Eur J Endocrinol. 2009;160(5):855–862.

60. Unsal T, et al. Genetic polymorphisms of FSHR, CYP17, CYP1A1, CAPN10, INSR, SERPINE1 genes in adolescent girls with polycystic ovary syndrome. J Assist Reprod Genet. 2009;26:205-216.

61. Ioannidis A, et al. Polymorphisms of the insulin receptor and the insulin receptor substrates genes in polycystic ovary syndrome: a Mendelian randomization meta-analysis. Mol Genet Metab. 2010;99(2):174–183.

62. Goodarzi MO, et al. Replication of association of a novel insulin receptor gene polymorphism with polycystic ovary syndrome. Fertil Steril. 2011;95(5):1736–1741. e1711.

63. Christopoulos P, et al. Genetic variants in TCF7L2 and KCNJ11 genes in a Greek population with polycystic ovary syndrome. Gynecol Endocrinol. 2008;24(9):486–490.

64. Tu X, et al. LEPR Gene Polymorphism and Plasma Soluble Leptin Receptor Levels are Associated with Polycystic Ovary Syndrome in Han Chinese Women. Per Med. 2017;14(4):299–307.

65. Liang J, et al. Associations of Leptin Receptor and Peroxisome Proliferator-Activated Receptor Gamma Polymorphisms with Polycystic Ovary Syndrome: A Meta-Analysis. Ann Nutr Metab. 2019;75(1):1–8.

66. Vanamala K, and Medithi S. Prospective role of leptin receptor gene and its polymorphisms on the onset of polycystic ovarian syndrome. Curr Womens Health Rev. 2024;20(2):120–133.

67. Sampurna K, et al. Genotype-Phenotype correlations of Leptin receptor gene in Polycystic ovarian disease (PCOS). IOSR J Biotechnol Biochem. 2018;4(1):20–27.

68. Dallel M, et al. Contrasting association of Leptin receptor polymorphisms and haplotypes with polycystic ovary syndrome in Bahraini and Tunisian women: a case– control study. Biosci Rep. 2021;41(1).

69. Ashrafi Mahabadi S, and Tafvizi F. Reduction of Soluble Leptin Receptor Levels in Women with Unexplained Infertility and the Effect of Leptin Receptor Gln223Arg Polymorphism on its Serum Level. J Gynaecol Obstet Cancer Res. 2020;5(4):149–158.

70. Lewandowski KC, et al. Familial partial lipodystrophy as differential diagnosis of polycystic ovary syndrome. Endokrynol Pol. 2015;66(6):550–554.

71. Gambineri A, et al. Monogenic polycystic ovary syndrome due to a mutation in the lamin A/C gene is sensitive to thiazolidinediones but not to metformin. Eur J Endocrinol. 2008;159(3):347–353.

72. Urbanek M, et al. The role of genetic variation in the lamin a/c gene in the etiology of polycystic ovary syndrome. J Clin Endocrinol Metab. 2009;94(7):2665–2669.

73. Maciel GAR, et al. LMNA Gene Variations in PCOS: A Persistent Genetic Clue. J Clin Endocrinol Metab. 2025:dgaf072.

74. Ewens KG, et al. FTO and MC4R Gene Variants Are Associated with Obesity in Polycystic Ovary Syndrome. PLoS One. 2011;6(1):e16390.

75. Tan S, et al. Large effects on body mass index and insulin resistance of fat mass and obesity associated gene (FTO) variants in patients with polycystic ovary syndrome (PCOS). BMC Med Genet. 2010;11(1):12.

76. Wu R, and Gragnoli C. The melanocortin receptor genes are linked to and associated with the risk of polycystic ovary syndrome in Italian families. J Ovarian Res. 2024;17(1):242.

77. Batarfi AA, et al. MC4R variants rs12970134 and rs17782313 are associated with obese polycystic ovary syndrome patients in the Western region of Saudi Arabia. BMC Med Genet. 2019;20(1):144.

78. Wang F, et al. Differential genes in adipocytes induced from polycystic and non-polycystic ovary syndrome-derived human embryonic stem cells. Syst Biol Reprod Med. 2014;60(3):136–142.

79. Li C, et al. In utero bisphenol A exposure disturbs germ cell cyst breakdown through the PI3k/Akt signaling pathway and BDNF expression. Ecotoxicol Environ Saf. 2023;259:115031.

80. Zhang Q, et al. Effects of brain-derived neurotrophic factor on oocyte maturation and embryonic development in a rat model of polycystic ovary syndrome. Reprod Fertil Dev. 2016;28(12):1904–1915.

81. Zhang Y, et al. Moderate Aerobic Exercise Regulates Follicular Dysfunction by Initiating Brain-Derived Neurotrophic Factor (BDNF)-Mediated Anti-Apoptotic Signaling Pathways in Polycystic Ovary Syndrome. J Clin Med. 2022;11(19):5584.

82. Schweighofer N, et al. Identification of Novel Intronic SNPs in Transporter Genes Associated with Metformin Side Effects. Genes. 2023;14(8):1609.

83. Højlund K, et al. Impaired Insulin-Stimulated Phosphorylation of Akt and AS160 in Skeletal Muscle of Women With Polycystic Ovary Syndrome Is Reversed by Pioglitazone Treatment. Diabetes. 2008;57(2):357–366.

84. Apperley L, et al. A rare case of congenital hyperinsulinism (CHI) due to dual genetic aetiology involving HNF4A and ABCC8. J Pediatr Endocrinol Metab. 2019;32(3):301–304.

85. Bohnen MS, et al. Loss-of-Function *ABCC8* Mutations in Pulmonary Arterial Hypertension. Circ Genom Precis Med. 2018;11(10):e002087.

86. Tatsi EB, et al. Next generation sequencing targeted gene panel in Greek MODY patients increases diagnostic accuracy. Pediatr Diabetes. 2020;21(1):28–39.

87. Bellanné-Chantelot C, et al. *ABCC8* and *KCNJ11* molecular spectrum of 109 patients with diazoxide-unresponsive congenital hyperinsulinism. J Med Genet. 2010;47(11):752–759.

88. Pinney SE, et al. Clinical characteristics and biochemical mechanisms of congenital hyperinsulinism associated with dominant K ATP channel mutations. J Clin Invest. 2008;118(8):2877–2886.

89. Martínez R, et al. Clinical and genetic characterization of congenital hyperinsulinism in Spain. Eur J Endocrinol. 2016;174(6):717–726.

90. Motte-Signoret E, et al. Glucocorticoid-Induced Hyperinsulinism in a Preterm Neonate with Inherited ABCC8 Variant. Metabolites. 2022;12(9):847.

91. Fernández–Marmiesse A, et al. Mutation spectra of ABCC8 gene in Spanish patients with Hyperinsulinism of Infancy (HI). Hum Mutat. 2006;27(2):214–214.

92. Schweiger B, et al. Reintroduction of Diazoxide after Diagnosis of Pulmonary Hypertension in a Patient with Transient Hyperinsulinism. J Child Sci. 2021;11(01):e80–e82.

93. Otonkoski T, et al. Noninvasive Diagnosis of Focal Hyperinsulinism of Infancy With [18F]-DOPA Positron Emission Tomography. Diabetes. 2006;55(1):13–18.

94. Ateş EA, et al. Genetic and Clinical Characterization of Patients with Maturity-Onset of Diabetes of the Young (MODY): Identification of Novel Variations. Balkan Med J. 2021;38(5):272–277.

95. Özdemir TR, et al. Targeted next generation sequencing in patients with maturity-onset diabetes of the young (MODY). J Pediatr Endocrinol Metab. 2018;31(12):1295–1304.

96. Riveline J-P, et al. Clinical and Metabolic Features of Adult-Onset Diabetes Caused by ABCC8 Mutations. Diabetes Care. 2012;35(2):248–251.

97. Li C, et al. Functional and Metabolomic Consequences of KATP Channel Inactivation in Human Islets. Diabetes. 2017;66(7):1901–1913.

98. De Franco E, et al. Update of variants identified in the pancreatic β-cell KATP channel genes KCNJ11 and ABCC8 in individuals with congenital hyperinsulinism and diabetes. Hum Mutat. 2020;41(5):884–905.

99. Lago-Docampo M, et al. Characterization of rare ABCC8 variants identified in Spanish pulmonary arterial hypertension patients. Sci Rep. 2020;10(1):15135.

100. Bennett JT, et al. Molecular genetic testing of patients with monogenic diabetes and hyperinsulinism. Mol Genet Metab. 2015;114(3):451–458.

101. Jahnavi S, et al. Novel ABCC8 (SUR1) gene mutations in Asian Indian children with congenital hyperinsulinemic hypoglycemia. Ann Hum Genet. 2014;78(5):311–319.

102. Nóvoa-Medina Y, et al. Congenital hyperinsulinism in Gran Canaria, Canary Isles. An Pediatr (Engl Ed). 2021;95(2):93–100.

103. Dallali H, et al. Genetic characterization of suspected MODY patients in Tunisia by targeted next-generation sequencing. Acta Diabetol. 2019;56(5):515–523.

104. Sun B, et al. [Novel MFN2, BSCL2 and LRSAM1 variants in a cohort of Chinese patients with Charcot-Marie-Tooth disease]. Zhonghua Nei Ke Za Zhi. 2022;61(8):901-907.

105. Jiang F, et al. Functional characterization of a novel heterozygous mutation in the glucokinase gene that causes mody2 in Chinese pedigrees. Front Endocrinol (Lausanne). 2021;12:803992.

106. Aykut A, et al. Analysis of the GCK gene in 79 MODY type 2 patients: A multicenter Turkish study, mutation profile and description of twenty novel mutations. Gene. 2018;641:186–189.

107. Capuano M, et al. Glucokinase (GCK) Mutations and Their Characterization in MODY2 Children of Southern Italy. PLoS One. 2012;7(6):e38906.

108. Yorifuji T, et al. Targeted gene panel analysis of Japanese patients with maturity-onset diabetes of the young-like diabetes mellitus: Roles of inactivating variants in the ABCC8 and insulin resistance genes. J Diabetes Investig. 2023;14(3):387–403.

109. Elashi AA, et al. The Genetic Spectrum of Maturity-Onset Diabetes of the Young (MODY) in Qatar, a Population-Based Study. Int J Mol Sci. 2023;24(1):130.

110. Yorifuji T, et al. Genetic basis of early-onset, maturity-onset diabetes of the young-like diabetes in Japan and features of patients without mutations in the major MODY genes: dominance of maternal inheritance. Pediatr Diabetes. 2018;19(7):1164–1172.

111. Matyka KA, et al. Genetic testing for maturity onset diabetes of the young in childhood hyperglycaemia. Arch Dis Child. 1998;78(6):552–554.

112. Delvecchio M, et al. Low Prevalence of HNF1A Mutations After Molecular Screening of Multiple MODY Genes in 58 Italian Families Recruited in the Pediatric or Adult Diabetes Clinic From a Single Italian Hospital. Diabetes Care. 2014;37(12):e258–e260.

113. Katashima R, et al. Identification of Novel GCK and HNF4α Gene Variants in Japanese Pediatric Patients with Onset of Diabetes before 17 Years of Age. J Diabetes Res. 2021;2021(1):7216339.

114. Estalella I, et al. Mutations in GCK and HNF-1α explain the majority of cases with clinical diagnosis of MODY in Spain. Clin Endocrinol (Oxf). 2007;67(4):538–546.

115. Borowiec M, et al. Novel glucokinase mutations in patients with monogenic diabetes – clinical outline of GCK-MD and potential for founder effect in Slavic population. Clin Genet. 2012;81(3):278–283.

116. Zhou Y, et al. MODY2 in Asia: analysis of GCK mutations and clinical characteristics. Endocr Connect. 2020;9(5):471–478.

117. Zhou Q, et al. Molecular and Clinical Profiles of Pediatric Monogenic Diabetes Subtypes: Comprehensive Genetic Analysis of 138 Patients. J Clin Endocrinol Metab. 2024.

118. Sagen JV, et al. Diagnostic screening of MODY2/GCK mutations in the Norwegian MODY Registry. Pediatr Diabetes. 2008;9(5):442–449.

119. Pruhova S, et al. Glucokinase diabetes in 103 families from a country-based study in the Czech Republic: geographically restricted distribution of two prevalent GCK mutations. Pediatr Diabetes. 2010;11(8):529–535.

120. Rees MG, et al. Correlation of rare coding variants in the gene encoding human glucokinase regulatory protein with phenotypic, cellular, and kinetic outcomes. J Clin Invest. 2012;122(1):205–217.

121. Jin J-L, et al. Intensive genetic analysis for Chinese patients with very high triglyceride levels: Relations of mutations to triglyceride levels and acute pancreatitis. eBioMedicine. 2018;38:171–177.

122. Tada H, et al. Clinical whole exome sequencing in severe hypertriglyceridemia. Clin Chim Acta. 2019;488:31–39.

123. Matsunaga A, et al. Variants of Lipid-Related Genes in Adult Japanese Patients with Severe Hypertriglyceridemia. J Atheroscler Thromb. 2020;27(12):1264–1277.

124. Shetty S, et al. Type 1 Hyperlipoproteinemia Due to Compound Heterozygous Rare Variants in GCKR. J Clin Endocrinol Metab. 2016;101(11):3884–3887.

125. O’Rahilly S, et al. Detection of Mutations in Insulin-Receptor Gene in NIDDM Patients by Analysis of Single-Stranded Conformation Polymorphisms. Diabetes. 1991;40(6):777–782.

126. Hashemian S, et al. Genotyping of ABCC8, KCNJ11, and HADH in Iranian Infants with Congenital Hyperinsulinism. Case Rep Endocrinol. 2021;2021(1):8826174.

127. Courbage S, et al. Implication of Heterozygous Variants in Genes of the Leptin– Melanocortin Pathway in Severe Obesity. J Clin Endocrinol Metab. 2021;106(10):2991–3006.

128. Hakim A. A Genomic Approach To Idiopathic Liver Disease In Adults. 2019.

129. Welling MS, et al. The Narrative of a Patient with Leptin Receptor Deficiency: Personalized Medicine for a Rare Genetic Obesity Disorder. Obes Facts. 2023;16(5):514–518.

130. Farooqi IS, et al. Clinical and molecular genetic spectrum of congenital deficiency of the leptin receptor. N Engl J Med. 2007;356(3):237–247.

131. Patni N, et al. Regional Body Fat Changes and Metabolic Complications in Children With, Lipodystrophy-Causing LMNA Variants. J Clin Endocrinol Metab. 2018;104(4):1099–1108.

132. Vasandani C, et al. Phenotypic Differences Among Familial Partial Lipodystrophy Due to LMNA or PPARG Variants. J Endocr Soc. 2022;6(12).

133. Youn G, et al. Autosomal recessive LMNA mutation causing restrictive dermopathy. Clin Genet. 2010;78(2):199–200.

134. Starke S, et al. Progeroid laminopathy with restrictive dermopathy-like features caused by an isodisomic LMNA mutation p. R435C. Aging (Albany NY). 2013;5(6):445.

135. Madej-Pilarczyk A, et al. Progeroid syndrome with scleroderma-like skin changes associated with homozygous R435C LMNA mutation. Am J Med Genet A. 2009;149(11):2387–2392.

136. Quijano-Roy S, et al. De novo LMNA mutations cause a new form of congenital muscular dystrophy. Ann Neurol. 2008;64(2):177–186.

137. Rodríguez Rondón AV, et al. MC4R Variants Modulate α-MSH and Setmelanotide Induced Cellular Signaling at Multiple Levels. J Clin Endocrinol Metab. 2024;109(10):2452–2466.

138. LUBRANO-BERTHELIER C, et al. Molecular Genetics of Human Obesity-Associated MC4R Mutations. Ann N Y Acad Sci. 2003;994(1):49–57.

139. Vaisse C, et al. Melanocortin-4 receptor mutations are a frequent and heterogeneous cause of morbid obesity. J Clin Invest. 2000;106(2):253–262.

140. Santos JL, et al. Obesity and eating behaviour in a three-generation chilean family with carriers of the Thr150Ile mutation in the melanocortin-4 receptor gene. J Physiol Biochem. 2008;64(3):205–210.

141. Vega JA, et al. Melanocortin-4 receptor gene variation is associated with eating behavior in Chilean adults. Ann Nutr Metab. 2016;68(1):35–41.

142. Ochoa MC, et al. A novel mutation Thr162Arg of the melanocortin 4 receptor gene in a Spanish children and adolescent population. Clin Endocrinol (Oxf). 2007;66(5):652–658.

143. Albuquerque D, et al. Novel Variants in the MC4R and LEPR Genes among Severely Obese Children from the Iberian Population. Ann Hum Genet. 2014;78(3):195–207.

144. Nishigori H, et al. Mutations in the small heterodimer partner gene are associated with mild obesity in Japanese subjects. Proc Natl Acad Sci. 2001;98(2):575–580.

145. Lee YS, et al. A POMC variant implicates β-melanocyte-stimulating hormone in the control of human energy balance. Cell Metab. 2006;3(2):135–140.

146. Challis BG, et al. A missense mutation disrupting a dibasic prohormone processing site in pro-opiomelanocortin (POMC) increases susceptibility to early-onset obesity through a novel molecular mechanism. Hum Mol Genet. 2002;11(17):1997–2004.

147. Miraglia del Giudice E, et al. Molecular screening of the proopiomelanocortin (POMC) gene in Italian obese children: report of three new mutations. Int J Obes. 2001;25(1):61-67.

148. Creemers JWM, et al. Mutations in the Amino-Terminal Region of Proopiomelanocortin (POMC) in Patients with Early-Onset Obesity Impair POMC Sorting to the Regulated Secretory Pathway. J Clin Endocrinol Metab. 2008;93(11):4494–4499.

149. Nordang GBN, et al. Next-generation sequencing of the monogenic obesity genes LEP, LEPR, MC4R, PCSK1 and POMC in a Norwegian cohort of patients with morbid obesity and normal weight controls. Mol Genet Metab. 2017;121(1):51-56.

150. Manco L, et al. Next-generation sequencing of 12 obesity genes in a Portuguese cohort of patients with overweight and obesity. Eur J Med Genet. 2023;66(4):104728.

151. Shabana, and Hasnain S. Prevalence of POMC R236G mutation in Pakistan. Obes Res Clin Pract. 2016;10:S110–S116.

152. Hosoe J, et al. Clinical usefulness of multigene screening with phenotype-driven bioinformatics analysis for the diagnosis of patients with monogenic diabetes or severe insulin resistance. Diabetes Res Clin Pract. 2020;169:108461.

